# Digitising point of care HIV test results to accurately measure, and improve performance towards, the UNAIDS 90-90-90 targets

**DOI:** 10.1101/19012302

**Authors:** Nisha Jacob, Brian Rice, Emma Kalk, Alexa Heekes, Jennie Morgan, James Hargreaves, Andrew Boulle

## Abstract

**Introduction:** High rates of pre-treatment loss to care among persons diagnosed with HIV persist. Linkage to care can be improved through active digitally-based surveillance. Currently, record-keeping for HIV diagnoses in South Africa is paper-based. Aggregated testing data are reported routinely, and only discordant findings result in a specimen being tested at a laboratory and digitised.

**Methods:** The Western Cape Province in South Africa has a Provincial Health Data Centre (PHDC) where person-level routine data are consolidated in a single database, leveraging the existence of a unique patient identifier. To facilitate improved surveillance, a pre-carbonated point-of-care test (PoCT) form was piloted, where one copy was routed to the centralised laboratory and digitised for PHDC inclusion. We evaluated the utility of the intervention using cross-sectional and retrospective cohort analyses, as well as comparisons with reported aggregate data.

**Results:** From May 2017 to June 2018, 11337 digitised point-of-care HIV testing records were linked to the PHDC. Overall, 96% of records in the aggregate dataset were digitised, with 97% linked to the PHDC. Of those tested, 79% were female (median age 27 years). Linkage demonstrated that 51.3% of patients testing HIV-positive were retesting. Of those truly newly diagnosed, 81% were linked to HIV care and 25% were initiated on antiretroviral therapy immediately.

**Conclusion:** Digitisation of PoCT results is feasible and provides individuated HIV testing data to assist in linkage to care and in differentiating newly diagnosed patients from positive patients retesting. Actionable and accurate data can improve the measurement of performance towards the UNAIDS 90-90-90 targets.

## Introduction

The last decade has seen marked changes in the management of the global HIV epidemic. The United Nation’s programme on HIV/AIDS (UNAIDS) set the 90-90-90 targets in 2015, aiming to diagnose 90% of those who are HIV infected, treat 90% of those diagnosed HIV-positive and virally suppress 90% of those receiving antiretroviral treatment [1]. These targets provide a framework for the universal test-and-treat principle introduced by the World Health Organization (WHO) in 2015 [2].

HIV diagnosis is the first step of several along the HIV treatment cascade towards viral suppression [1]. That many people do not immediately proceed beyond this first step reflects a system failure; high rates of pre-treatment loss-to-care among persons diagnosed HIV-positive have been reported in sub-Saharan Africa [3][4][5]. If we are to meet the 90-90-90 targets it is essential that we improve, and accurately measure, linkage to HIV care for those newly diagnosed with HIV.

In South Africa, as with many countries in the region, HIV testing is based on a point-of-care test (PoCT) algorithm; only discordant findings result in a specimen being sent to a laboratory. This is against a backdrop of what is otherwise a comprehensive national laboratory information system (LIS) in which all laboratory test results performed anywhere in the country in the public sector are accessible on a central LIS, and can contribute to surveillance activities [6]. The move towards universal access to treatment, and high targets for identifying those infected with HIV, requires that the treatment cascade is monitored from the point of HIV testing. At testing facilities, PoCT HIV results are currently entered manually into a paper-based register, with aggregated data then submitted to the District Health Information System on a monthly basis.

The Western Cape Province has recently developed a Provincial Health Data Centre (PHDC) in which all person-level hospital and clinic administrative, laboratory, and pharmacy records, and data from disease information systems for HIV and TB, are consolidated in a single environment, leveraging the existence of a unique patient identifier which is used as the folder number in all health facilities in the public sector [7]. Within this environment, disease-specific patient cascades (virtual cohorts), such as the HIV treatment cascade, are in development using specific markers of care. In the HIV treatment cascade, electronic evidence of having HIV is based on laboratory markers (where PoCT is discordant or contra-indicated, or from other laboratory tests often conducted shortly after diagnosis, such as CD4 tests and viral load) as well as ICD-10 coding, for which the poor completeness and quality in the public sector is known [8–10]. The HIV cascade includes treatment information, such as initiation dates, regimen details, last dispensing dates, as well as information on viral suppression.

Recently, considerable media attention was given to a proposed PEPFAR funding cut for South Africa due to apparent poor performance of South African HIV programmes [11]. Although this proposed funding cut was later reversed, it was widely appreciated that improved performance could be supported through digital solutions which include routine electronic monitoring of all persons newly diagnosed with HIV and their subsequent care.

To facilitate improved HIV surveillance, the Department of Health in the Western Cape implemented a pilot intervention to digitise PoCT HIV test results with support from the laboratory transport mechanism and information system. PoCT results were thus available electronically at a population *and* an individual level. In this study, we evaluated the feasibility and utility of this pilot intervention at a local Community Health Centre (CHC), in meeting reporting requirements, and more importantly providing a province-wide individuated actionable reporting mechanism for assessing and improving linkage to care of clients newly diagnosed with HIV.

## Methods

The intervention was evaluated by applying cross-sectional and retrospective cohort analyses, as well as comparisons with reported aggregate data. The study population included all patients accessing HIV Testing Services (HTS) at Gugulethu CHC between May 2017 to June 2018. This included patients attending HTS as part of routine antenatal care, patients referred for HTS and those self-presenting for HTS. No formal recruitment was required as existing records in routine databases housed by the Department of Health were used for the evaluation.

At the facility, trained HIV counsellors obtained informed consent and performed the rapid PoCT HIV assays as per National protocol [12]. Carbonated copies of standard HTS forms completed at the facility were transported via routine systems to a central point for data capturing and inclusion in the LIS and PHDC. From May to December 2017 (Phase I) data was entered by a dedicated capturer located at the Provincial Department of Health. From January to June 2018 (Phase II), capturing was undertaken by data clerks at the National Health Laboratory Service (NHLS). Elements documented on the paper forms and digitised included patient identifiers, demographic data, testing consent, self-reported previous HIV test result, reason for accessing HTS and results of the rapid HIV PoCT. HIV status was calculated using the National Department of Health HIV testing algorithm [12].

Data were analysed using Microsoft Excel and Stata 14 (Stata Corporation, College Station, Texas, USA). The PHDC provided de-identified linked data for the analyses at study closure.

Patients attending HTS were described using appropriate descriptive statistics. Data were linked to the Patient Master Index and existing HIV cascade of the PHDC using patient folder numbers. Where folder numbers were unavailable or illegible, linkage was attempted using combinations of other patient identifiers such as name, surname, civil identifier and date of birth. All patients with evidence of HIV prior to PoCT were termed “cascade known HIV-positive”. Retesting was determined using self-reported HIV status and cascade status. Patients who self-reported a previous positive HIV test result and tested positive again during the pilot PoCT were termed “self-reported retest HIV-positive”. Patients with prior evidence of HIV but self-reported a previous negative or unknown HIV status and had a calculated HIV-positive test outcome were termed “unreported retested HIV-positive”. Patients with no prior evidence of HIV, who had a calculated HIV-positive test outcome were termed “true newly diagnosed HIV-positive”.

To assess linkage to care, cohort analyses were conducted for all patients who tested HIV-positive using laboratory and pharmacy indicators of HIV care, including viral load and antiretroviral treatment (ART). ART uptake was calculated for patients who were not immediately initiated on treatment following a positive HIV PoCT result.

Multivariate logistic regression was used to explore associations with retesting in patients previously diagnosed with HIV. Variables were selected a priori and included in the model to address the effects of confounding. Ethical approval for the study was obtained from the University of Cape Town Human Research Ethics Committee (HREC 198/2018).

## Results

From May 2017 to June 2018, 11 337 HIV counselling and testing forms were captured. Over this period, all digitised HIV PoCT records were linked to the PHDC to determine the contribution of the testing data to earlier ascertainment of HIV status and linkage to care. Table 1 shows the key descriptive characteristics of patients tested during the pilot intervention. Most patients were female (79.3%), with many presenting for antenatal care (39.8%). Only 13.4% of patients reported not having ever tested previously.

**Table 1:**
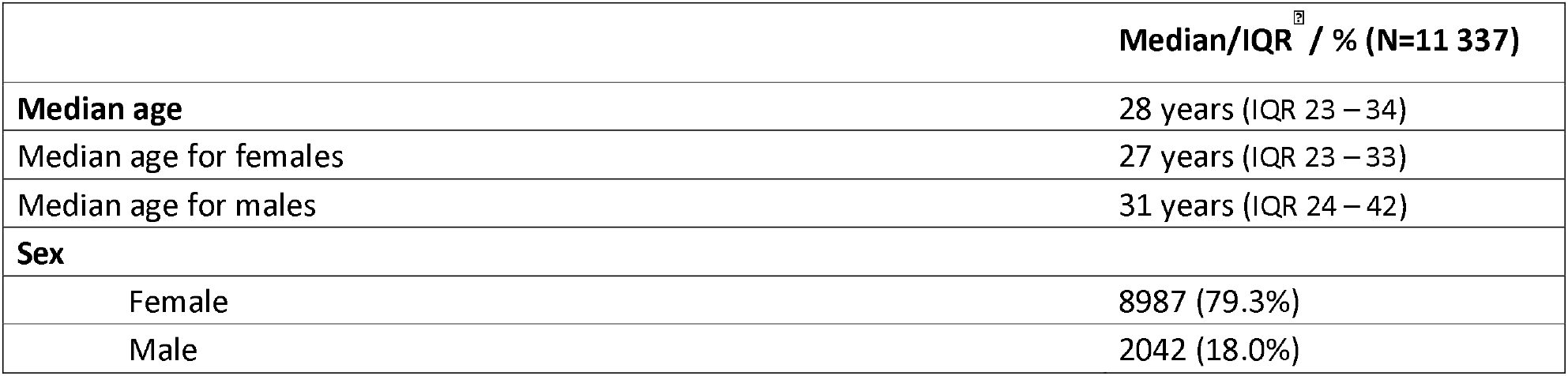
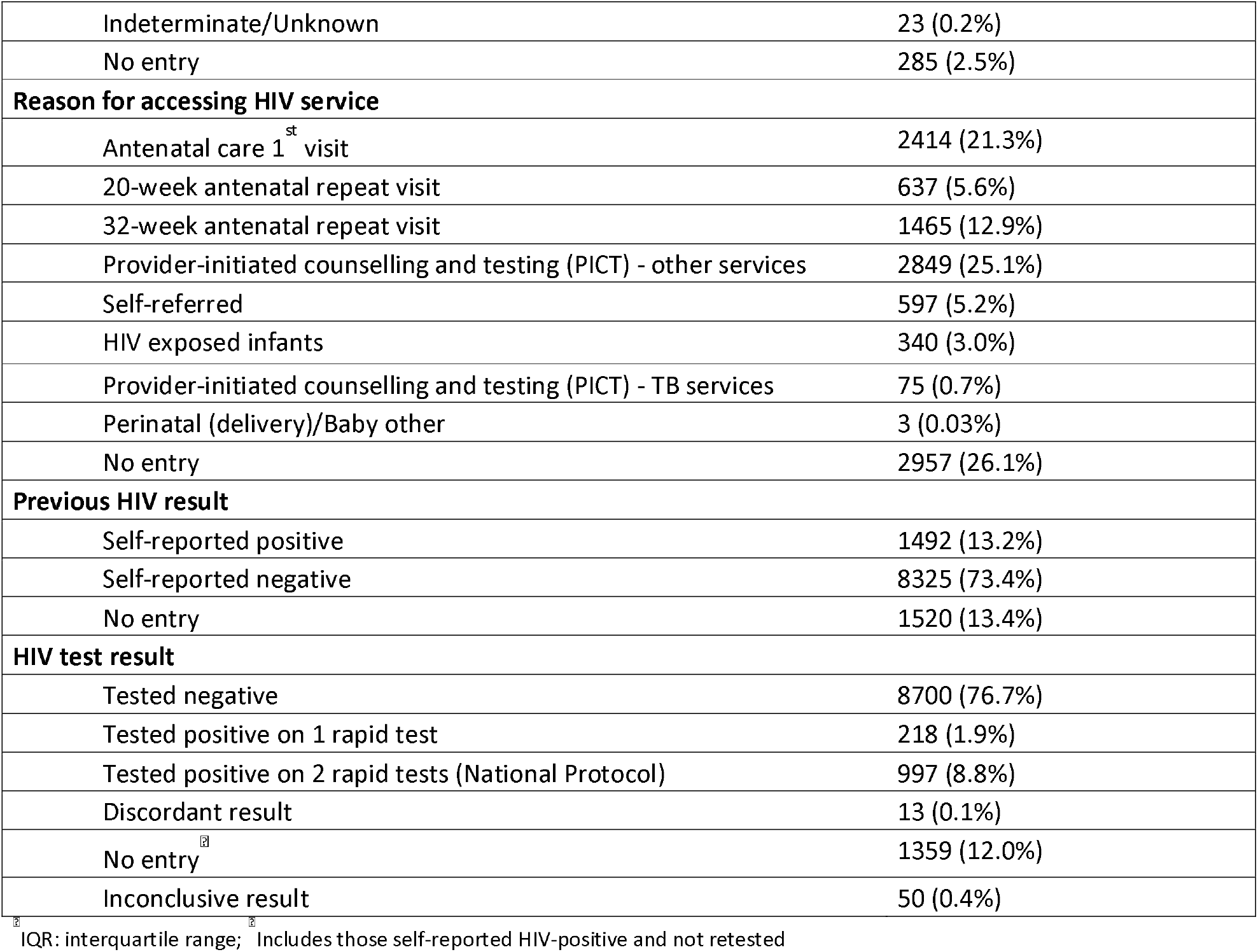
Characteristics of patients attending HTS

### Digitisation of records

During the study period, 11564 PoCT entries were tallied from paper registers as part of routine aggregate reporting over 13 completed months (May 2017 – May 2018). Therefore, 96.0% (11119/11564) of entries in the aggregate dataset were individually digitised as part of this project.

Overall, 97.0% of records could be linked to the Patient Master Index. Unlinked records could not be definitively linked to a single patient with the available identifiers.

### HIV status: PoCT result, self-reporting and re-testing

Of the 11337 patients, 76.7% (8700) tested HIV negative, with 10.7% (1215) testing positive. The remaining 12.5% (1422) did not have a conclusive PoCT outcome recorded. The overall prevalence of HIV among those attending HTS, was 21.2% (2399), almost half of whom (1132) self-reported a positive HIV status and did not retest.

Linkage of the apparent 907 newly diagnosed HIV-positive patients to the HIV cascade indicated that over a third (34.7%; 315) were already known to the cascade as HIV-positive, supporting an established diagnosis of HIV in the past. These 315 patients were therefore retesting following a previous positive test results, having not self-reported a positive HIV status. Of those who retested with reporting their HIV-positive status, 46.7% (147/315) had commenced ART prior to their latest PoCT. Overall, 51.3% (623/1215) of patients who tested HIV-positive on at least one rapid test had previously been diagnosed (Fig 1).

**Fig 1:**
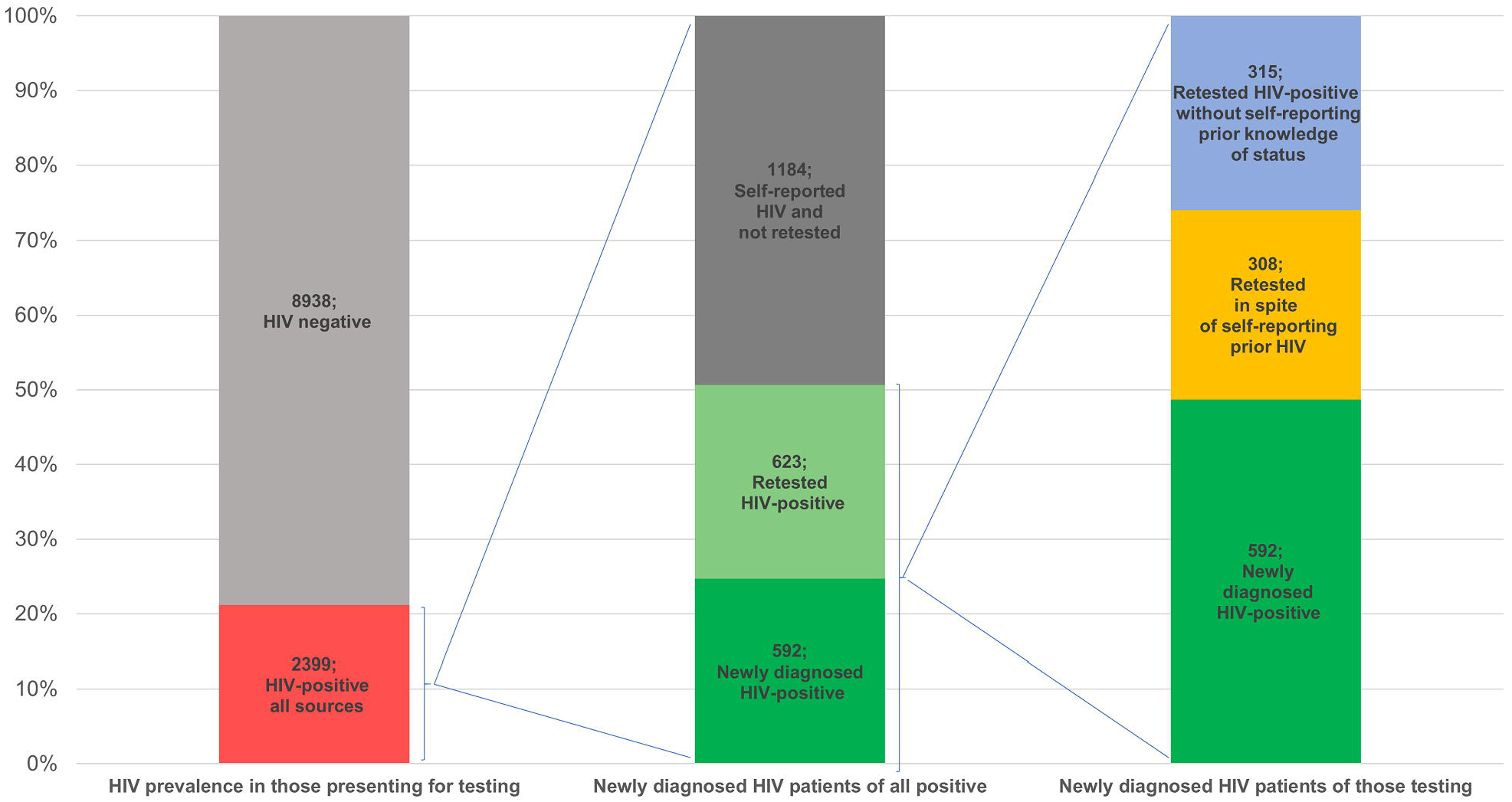
HIV positivity and yield by testing and prior testing status.

Of patients who self-reported a positive HIV status, 88.2% (1316/1492) were already known to the PHDC HIV cascade as HIV-positive. Based on available laboratory and pharmacy evidence, an additional 8.3% (125) entered the HIV cascade after their HIV PoCT during the pilot intervention. The remaining 3.4% (51) did not have any prior or subsequent evidence of HIV and were thus not known to the HIV cascade. Of persons self-reporting a positive HIV status, 24.1% (360/1492) retested (self-reported retest). Fig 2 demonstrates the utility of digitisation and individual data linkage to the HIV cascade compared to information garnered from aggregate systems in isolation. Although 50 patients retested HIV negative, having self-reported HIV-positive status, linkage to the HIV cascade shows that 38 of these patients had previous evidence of HIV diagnosis in the HIV cascade.

**Fig 2:**
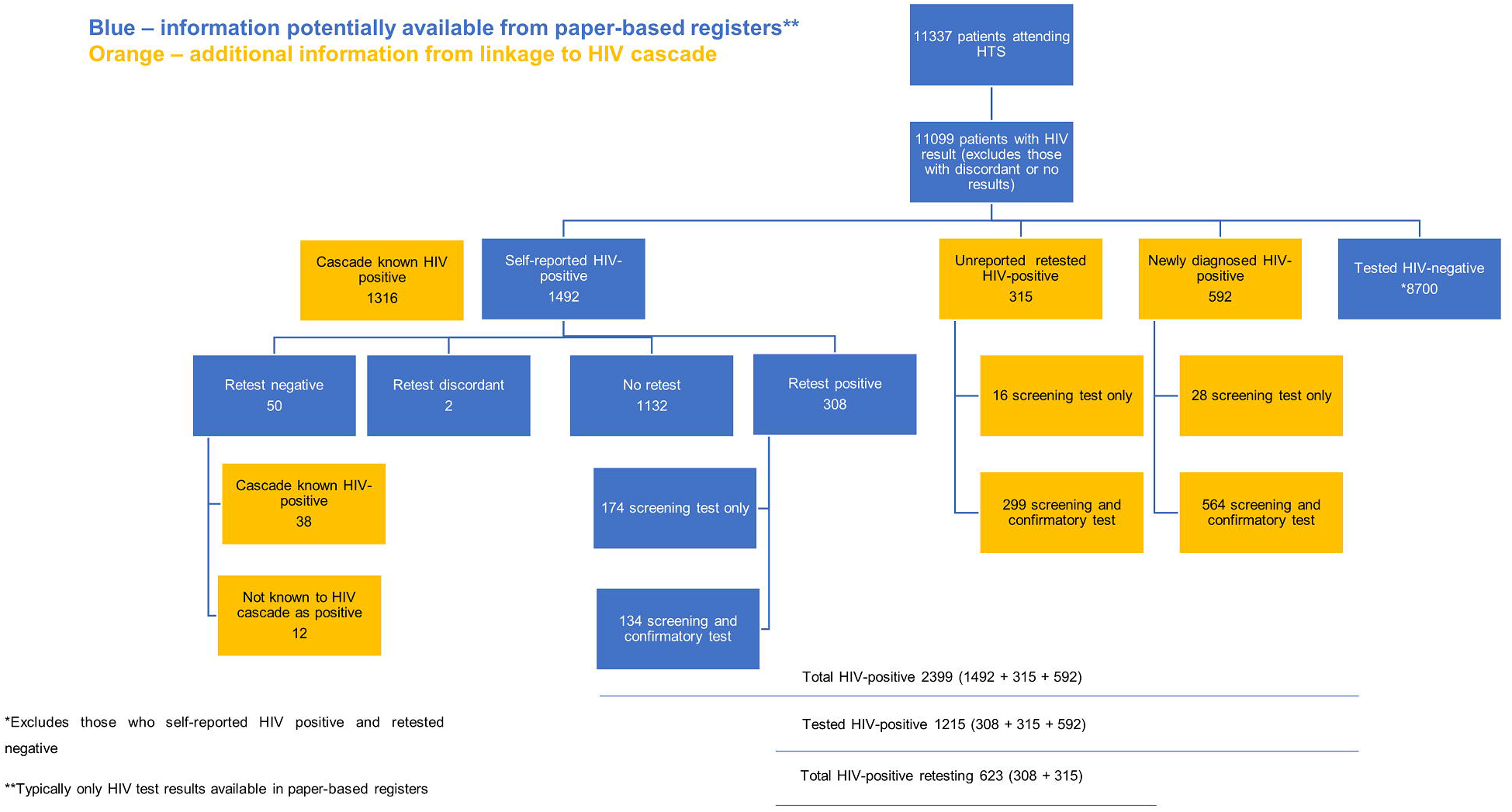
Contribution of HIV cascade data linkages to testing information.

### Factors associated with re-testing in patients with known HIV

Apparent bivariate associations of older age and women with retesting in spite of HIV-positive status having been ascertained previously were attenuated through adjusting for the reason for testing. In the adjusted model, reason for attending HTS services was highly associated with retesting (Table 2). For example, those attending for provider-initiated counselling and testing (PICT) from TB services were 10.6 (95% CI 2.8-39.7) times more likely to retest compared to those attending initial antenatal visits. The association between younger age and retesting also persisted.

**Table 2:** Factors associated with retesting in patients already diagnosed with HIV

| Variable | Unadjusted |  | Adjusted |  |
| --- | --- | --- | --- | --- |
|  | OR (95% CI) | p-value | OR (95% CI) | p-value |
| **Sex (reference female)** | 2.45 (1.91 – 3.15) | <0.001 | 1.50 (0.85 – 2.66) | 0.163 |
| **Age category (reference 25-49 years)** |  |  |  |  |
| 15 – 24 years | 1.39 (1.03 - 1.88) | 0.029 | 1.54 (1.10 – 2.16) | 0.012 |
| 25-49 years | 1.00 (ref) | 0.003 | 1.00 (ref) | 0.611 |
| >50 years | 4.17 (1.63 – 10.68) |  | 0.77 (0.29 – 2.08) |  |
| **ART status (reference prior ART)** |  |  |  |  |
| Prior ART | 1.00 (ref) | 0.022 | 1.00 (ref) | 0.075 |
| Current ART | 0.59 (0.37 – 0.93) | 0.041 | 1.64 (0.95 – 2.84) | 0.199 |
| No ART | 1.27 (0.77 – 2.10) |  | 1.47 (0.82 – 2.63) |  |
| **Reason for accessing service (reference Antenatal 1<sup>st</sup> visit)** |  |  |  |  |
| Antenatal 1<sup>st</sup> visit | 1.00 (ref) | <0.001 | 1.00 (ref) | <0.001 |
| Antenatal 1<sup>st</sup> visit | 0.09 (0.03 – 0.25) | 0.111 | 0.09 (0.03 – 0.25) | 0.092 |
| 20-week antenatal repeat visit | 0.72 (0.48 – 1.08) | <0.001 | 0.70 (0.47 – 1.06) | <0.001 |
| 32-week antenatal repeat visit | 10.54 (2.87 – 38.74) | <0.001 | 10.67 (2.86 – 39.73) | <0.001 |
| PICT TB services | 6.81 (5.03 – 9.24) | <0.001 | 7.07 (4.93 – 10.13) | <0.001 |
| PICT other services | 7.51 (3.23 – 17.47) | 0.111 | 7.71 (3.16 – 18.80) | 0.090 |
| Self-referred | 0.62 (0.35 – 1.12) |  | 0.60 (0.34 – 1.08) |  |
| HIV exposed infants |  |  |  |  |

### Linkage to care and ART

Of patients confirmed as being newly diagnosed HIV-positive (as per linked HIV cascade data), 81.3% (481/592) had evidence of further HIV care at the time of analysis, including 58.1% who had started ART (Fig 3). Of those who were retesting HIV-positive, 71.4% had already started ART previously, and a further 13.6% subsequently started ART (Fig 3).

**Fig 3:**
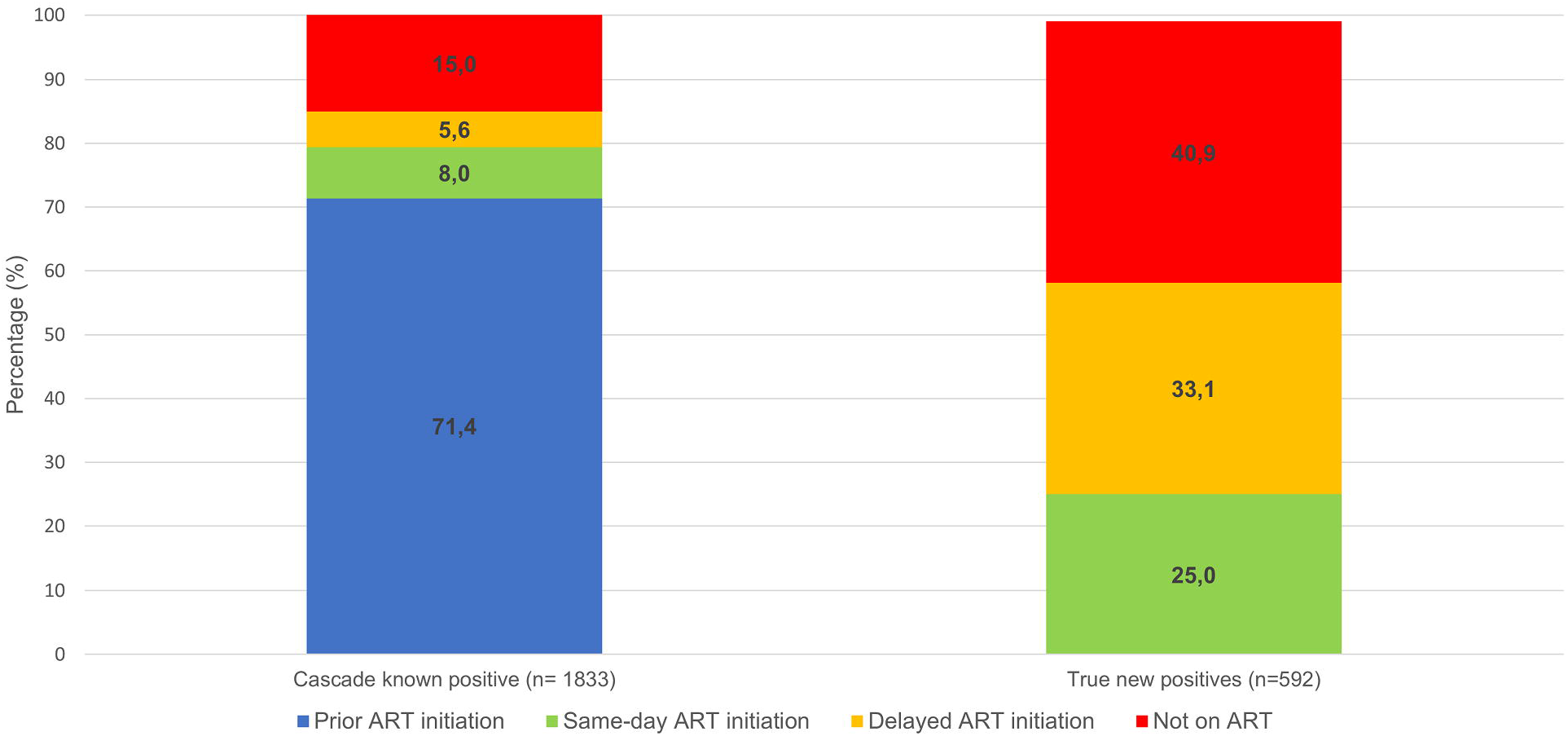
Antiretroviral (ART) status among cascade known positive and true new positive patients.

## Discussion

This study illustrates the important contribution of HIV PoCT data to earlier digital ascertainment of HIV status and linkage to care. We show that digitisation of PoCT records is feasible, having captured electronically 96.0% of expected records. Linkage of HIV PoCT data using the unique identifier further allows discernment of newly diagnosed HIV-positive patients from those who are retesting. We found that half of patients testing HIV-positive on at least one rapid test were retesting following a previous HIV diagnosis, reducing to a third if excluding those who retested positive in spite of indicating that they already knew they had HIV.

### Completeness of ascertainment and linkage of records

Linkage was close to 100% indicating that most patients who had attended HTS during the pilot were registered with public health care services. The majority of patients who self-reported a positive HIV status were known to the HIV cascade, validating the completeness of the cascade within the PHDC. Since HIV PoCT are not routinely digitised, and hence not part of the electronic PHDC HIV cascade, the 176 patients who self-reported HIV-positive in the pilot but did not appear in the cascade, most likely did not engage in further HIV-specific care following an earlier positive PoCT. Overall, 8.4% of patients who self-reported a positive HIV status only entered the HIV cascade following PoCT during the pilot study, when further HIV-specific management was initiated. This pre-treatment loss to care is well-documented throughout sub-Saharan Africa but notably challenging to quantify in the absence of robust health information systems [3][4].

The most recent population-level HIV survey in South Africa showed an HIV prevalence of 12.6% (95% CI 9.7 – 16.1) in the Western Cape province [13]. According to the most recent antenatal HIV prevalence survey, the Western Cape province HIV prevalence was 20.8% [14]. In this study, 10.7% of patients tested HIV-positive on at least one rapid test, although this includes retesting and excludes those who elected not to retest following a self-reported previous HIV-positive result. The overall prevalence of HIV among those presenting for HTS was 21.2%, including those who self-reported a previous positive result. This finding is similar to the overall provincial antenatal HIV prevalence and is expected as antenatal patients comprise a large proportion of the study population.

### Retesting in patients already known to have HIV

Retesting among patients with a previous positive HIV diagnosis but who self-reported a negative or unknown status was an important finding that is not quantifiable in the absence of PoCT digitisation. Paper-based records are limited in revealing this phenomenon due to the movement of patients between different public health facilities. We identified known HIV-positive patients as being a substantial proportion of those testing, and additionally identified a further proportion who were retesting without revealing their HIV status. Half of the patients who knew they had HIV and who retested did not declare a previous HIV-positive result and hence were misclassified as new HIV-positive diagnoses in aggregate data. In practice, all retesting is likely to be misclassified in aggregate data due to challenges in tallying data across multiple columns in paper-based registers. Such misclassification has significant implications for South Africa’s approach to measuring progress towards the UNAIDS 90-90-90 goals [1]. Considerable resources have been committed to expanding HIV testing and counselling programmes to achieve the first 90. Our results suggest a third (34.8%) of HIV-infected patients identified by programmes may already be aware of their status. Furthermore, of these patients, almost a half (46.7%) had evidence of prior ART. Attention thus needs to be directed at retaining such individuals in care and on ART (the second and third 90 targets).

### Understanding retesting

There is a paucity of literature exploring reasons for retesting after a positive HIV result, particularly among those who are already initiated on ART. Since all pregnant patients are referred to the HIV testing counsellors regardless of HIV or ART status, the opportunity for reaffirmation of diagnosis may have prompted retesting, however further research may be needed to better understand reasons for retesting more broadly.

Various factors may be associated with non-disclosure of HIV-positive status to healthcare professionals. Most studies have explored non-disclosure of HIV status to family and friends [15][16]. One study indicates that non-disclosure to health professionals is as high as 40% in certain settings [15]. Societal stigma, feelings of embarrassment, and denial may all contribute to non-disclosure and seeking repeat testing [3,15,17]. Another reason, less explored in the literature, is patient understanding of a diagnosis of HIV. A survey conducted in South Africa, showed that only 91% of patients with a previous HIV test were aware of their results [18]. Whether an awareness of a positive result translates to understanding the diagnosis is not well defined. While there are several studies that demonstrate the link between health literacy and health outcomes, most focus on print literacy [19,20]. In the South African setting, patients may be more reliant on oral literacy which may be limited by language barriers between patients and healthcare professionals, rushed consultations and long waiting times. Prior to implementation of the universal test-and-treat strategy in 2015, patients were not eligible for ART until certain criteria were met. This may have led to false perceptions that patients who tested positive for HIV but were not treated, and remained asymptomatic, did not have the infection. These factors may have resulted in apparent non-disclosure of HIV status and subsequent retesting, although further research is imperative to better understand this phenomenon.

Those who were referred for PoCT by healthcare workers in other CHC services were more likely to be retesting than those receiving PoCT as part of antenatal care. Younger patients were more likely to retest than older patients. Reasons for these associations require more detailed analyses with consideration for confounding factors such as educational status, socio-economic status, and family support. Linkage of PoCT results to a consolidated individuated data environment contributes to uncovering such phenomena and the ensuing bias introduced when interpreting HIV testing data.

### Linkage to care and treatment in those testing positive

Longitudinal analyses of individual patient data can assess implementation of HIV care guidelines, in this case, universal test-and-treat. Of those newly diagnosed with HIV, more than half (58.1%) were initiated on ART at study closure, while only a quarter (25%) initiated ART on the day of testing. Our results indicate that the universal test-and-treat strategy has not been fully implemented. Further research is needed to better understand why 41% of newly diagnosed HIV-positive patients had not initiated ART. Although reasons for non-initiation may not be explicit in data available on the PHDC, alerting systems can be developed to flag patients who have not engaged in appropriate HIV care at future consultations or to find and encourage patients to return to HIV care for further management.

## Limitations

There are various pitfalls of using routine data for monitoring and evaluation [21,22], particularly data accuracy and completeness. Since PoCT data are entered as routine clerical tasks in settings where staff have high patient loads, the risk of inaccuracies is high. The 50 patients who self-reported HIV-positive but retested negative likely represent capturing errors of the result on the HTS form, as three quarters of these patients had evidence of HIV in the cascade. However, further studies are warranted to explore whether these may represent patients who had a previous false positive HIV PoCT. Furthermore, high staff turnover among HIV counsellors and nursing staff may result in inadequately trained staff members conducting tests and completing forms, resulting in erroneous entries and incomplete data. The use of a single rapid test for many retesting HIV-positive patients was most likely for reaffirmation of diagnosis, hence the deviation from the National HIV testing protocol. However, there were patients without prior of evidence of HIV who had a single positive test or results entered in incorrect fields, suggesting training gaps. By uncovering such training gaps, digitisation may prompt targeted training interventions, thereby strengthening clinical governance. Limited staff involvement and understanding of data use may also contribute to de-prioritisation of data capturing in the face of more pressing clinical concerns [22]. The quality of PoCT and the validity of the results may be called to question as facility staff are not laboratory-trained and may not be able to adhere to strict quality control criteria [23]. Since HIV PoCT is the standard of care for HIV diagnosis and referral, digitisation of available results will nonetheless add value to the HIV cascade through earlier ascertainment of HIV status and linkage to care. Predictors for retesting could not be fully evaluated in this study, as variables were limited to routinely captured HTS forms. Data on other confounding factors were thus not collected and adjusted for.

## Conclusions

This study clearly demonstrates the important contribution of digitised PoCT data, within a robust PHDC system, to better evaluate the HIV treatment cascade at both a population and an individual level. Improved estimation of the magnitude of retesting may have substantial implications for assessing interventions to achieve 90-90-90 targets. Considerable resources are channelled into achieving the first 90 through wide-scale HIV testing. However, without quantifying the proportion of HIV-infected patients who are retesting, this target cannot be appropriately evaluated. Furthermore, the findings of this study demonstrate the contribution of PoCT in evaluating the second 90 using routine data within the PHDC, and differentiating those who need linkage to care following a new diagnosis, from those who need retention and re-engagement on ART.

While this study evaluated the population-level utility of PoCT, individual utility may also be demonstrated. Health care professional access to PoCT results along with other laboratory data, may enhance clinical consultations by improving efficiency during patient history-taking and avoiding duplication of tests, which are uncomfortable and invasive to patients as well as costly to the health system. Timely access to PoCT data and subsequent care access may provide actionable opportunities to follow-up individual patients who have not linked to care and may enhance operational management by facilitating improved monitoring of testing services. It is recommended that quantitative and qualitative studies are conducted to better evaluate these other perceived benefits of PoCT result digitisation.

## Data Availability

Data are not made available as they are unconsented, de-identified routine administrative data housed in the Western Cape Provincial Health Data Centre.

## Authorship

All authors have read and approved the final manuscript. NJ, AB, BR and JH designed the research study. NJ conducted the research. NJ and AB wrote the manuscript. BR and JH made significant intellectual contributions to the manuscript preparation. EK and JM provided practical support for the research and contributed to the manuscript preparation. AH provided technical support in data linkage and analyses.

## Acknowledgements

The authors gratefully acknowledge the contributions of various collaborators, including: Linda Hlwaya, Ezethu Sebezo, Nasima Mohamed, Igsaan Noordien, Izak de Villiers, Helena Vreede, Nontobeko Tena-Coki, Lunga Makamba, the National Health Laboratory Services, the staff, counsellors and patients at Gugulethu Community Health Centre and the Measurement and Surveillance of HIV Epidemics (MeSH) consortium.

